# Rapid implementation of a cohort for the study of post-acute sequelae of SARS-CoV-2 infection/COVID-19

**DOI:** 10.1101/2021.03.11.21252311

**Authors:** Michael J. Peluso, J. Daniel Kelly, Scott Lu, Sarah A. Goldberg, Michelle C. Davidson, Sujata Mathur, Matthew S. Durstenfeld, Matthew A. Spinelli, Rebecca Hoh, Viva Tai, Emily A. Fehrman, Leonel Torres, Yanel Hernandez, Meghann C. Williams, Mireya I. Arreguin, Jennifer A. Bautista, Lynn H. Ngo, Monika Deswal, Sadie E. Munter, Enrique O. Martinez, Khamal A. Anglin, Mariela D. Romero, Jacqueline Tavs, Paulina R. Rugart, Jessica Y. Chen, Hannah M. Sans, Victoria W. Murray, Payton K. Ellis, Kevin C. Donohue, Jonathan A. Massachi, Jacob O. Weiss, Irum Mehdi, Jesus Pineda-Ramirez, Alex F. Tang, Megan Wenger, Melissa Assenzio, Yan Yuan, Melissa Krone, Rachel L. Rutishauser, Isabel Rodriguez-Barraquer, Bryan Greenhouse, John A. Sauceda, Monica Gandhi, Priscilla Y. Hsue, Timothy J. Henrich, Steven G. Deeks, Jeffrey N. Martin

## Abstract

**BACKGROUND:** As the coronavirus disease 2019 (COVID-19) pandemic continues and millions remain vulnerable to infection with severe acute respiratory syndrome-coronavirus-2 (SARS-CoV-2), attention has turned to characterizing post-acute sequelae of SARS-CoV-2 infection (PASC).

**METHODS:** From April 21 to December 31, 2020, we assembled a cohort of consecutive volunteers who a) had documented history of SARS-CoV-2 RNA-positivity; b) were ≥ 2 weeks past onset of COVID-19 symptoms or, if asymptomatic, first test for SARS-CoV-2; and c) were able to travel to our site in San Francisco. Participants learned about the study by being identified on medical center-based registries and being notified or by responding to advertisements. At 4-month intervals, we asked participants about physical symptoms that were new or worse compared to the period prior to COVID-19, mental health symptoms and quality of life. We described 4 time periods: 1) acute illness (0-3 weeks), 2) early recovery (3-10 weeks), 3) late recovery 1 (12-20 weeks), and 4) late recovery 2 (28-36 weeks). Blood and oral specimens were collected at each visit.

**RESULTS:** We have, to date, enrolled 179 adults. During acute SARS-CoV-2 infection, 10 had been asymptomatic, 125 symptomatic but not hospitalized, and 44 symptomatic and hospitalized. In the acute phase, the most common symptoms were fatigue, fever, myalgia, cough and anosmia/dysgeusia. During the post-acute phase, fatigue, shortness of breath, concentration problems, headaches, trouble sleeping and anosmia/dysgeusia were the most commonly reported symptoms, but a variety of others were endorsed by at least some participants. Some experienced symptoms of depression, anxiety, and post-traumatic stress, as well as difficulties with ambulation and performance of usual activities. The median visual analogue scale value rating of general health was lower at 4 and 8 months (80, interquartile range [IQR]: 70-90; and 80, IQR 75-90) compared to prior to COVID-19 (85; IQR 75-90). Biospecimens were collected at nearly 600 participant-visits.

**CONCLUSION:** Among a cohort of participants enrolled in the post-acute phase of SARS-CoV-2 infection, we found many with persistent physical symptoms through 8 months following onset of COVID-19 with an impact on self-rated overall health. The presence of participants with and without symptoms and ample biological specimens will facilitate study of PASC pathogenesis. Similar evaluations in a population-representative sample will be needed to estimate the population-level prevalence of PASC.

## BACKGROUND

Coronavirus disease 2019 (COVID-19), the condition caused by severe acute respiratory syndrome-coronavirus-2 (SARS-CoV-2), was initially characterized as a time-limited illness [1–3]. Patients were believed to either succumb to COVID-19 or return to their usual health. Subsequently, anecdotal reports emerged stating that while recovery from the symptoms typically associated with an acute infection (e.g., fever and chills) is near uniform, some individuals complain of persistent symptoms (e.g., fatigue and pain) well after the period of acute SARS-CoV-2 infection [4–6]. These patients gave rise to the colloquial terms “long haulers” and “long COVID” [7,8]. Formal scientific investigation of what is clinically known as post-acute sequelae of SARS-CoV-2 infection (PASC) has just begun and has been useful to establish the frequency of the condition beyond anecdote and to demonstrate its geographic and sociodemographic diversity. These early studies have, however, been limited by studying populations who had not all been confirmed to have SARS-CoV-2 [9], were enriched with patients who were hospitalized and may thus be more indicative of the effects of hospitalization than COVID-19 [10,11], or had short follow-up [12]. Furthermore, the pathogenesis of PASC is entirely unknown. Some studies have detected biologic abnormalities but have not yet linked them to clinical phenotypes [13–18]. As millions of individuals worldwide continue to become infected with SARS-CoV-2, the public health implications of PASC and the need to uncover interventions to prevent or treat it are self-evident.

To rapidly gain insights into the period following acute SARS-CoV-2 infection, we urgently established a cohort dedicated to the study of PASC. We intentionally sought to enroll a large number of patients with RNA-confirmed SARS-CoV-2 infection recovering from a wide spectrum of acute disease manifestations, and we supplemented broad clinical phenotyping with ample biological specimen collection for pathogenesis-focused studies. Herein, we describe the spectrum of physical and mental health symptoms and quality of life through 8 months of observation following onset of acute symptoms as well as the resources available for collaborative research.

## METHODS

### Ethics Statement

The institutional review board of the University of California, San Francisco, approved this study. All participants provided written informed consent.

### Overall Design

We enrolled consecutive volunteers who were at least two weeks past their onset of COVID-19 symptoms or, if no symptoms, first positive diagnostic test for SARS-CoV-2 and who responded to notification and advertisements regarding the long-term consequences of COVID-19. Participants underwent comprehensive questionnaire-based evaluation and biological specimen collection at their initial visit and at approximately 4-month intervals thereafter.

### Participants

Beginning in April 2020, any individual age 18 years or older with documentation (either paper or electronic record) of prior SARS-CoV-2 RNA detected on a nucleic acid amplification test and ability to travel to our research site in San Francisco was eligible to participate. Minimal duration of time following symptom onset (or first positive RNA detection) depended on local infection control guidelines, starting with a minimum of 28 days and subsequently shortening to 14 days. Participants were recruited through clinician referral, mailings to consecutive patients testing positive at University of California, San Francisco- or San Francisco General Hospital-affiliated testing sites, and response to medical center paper postings, websites (including www.liincstudy.org and www.clinicaltrials.gov), and web-based advertisements. We also developed close partnerships with local studies of acute SARS-CoV-2 infection, offering long-term follow-up once the initial research period was complete. We sought diverse representation in terms of acute SARS-CoV-2 disease manifestations. Although quotas were not set, given that most acute SARS-CoV-2 infection does not result in severe disease or hospitalization, most recruitment resources were dedicated to attracting persons who had not been hospitalized, including those who had been asymptomatic.

### Processes

Once identified, participants deemed eligible by a phone interview were examined in person at the research center. Each participant had to pass a medical center-mandated COVID-19 symptom screening prior to entering the building; numbers of participants per day were limited by ambient workplace density reduction measures. Participants were administered structured questionnaires, asked to provide whole unstimulated saliva and/or a swab of gingival crevice fluid, and had peripheral blood collected, which was stored as serum, plasma, and cryopreserved peripheral blood mononuclear cells (PBMCs). Following the initial visit, participants were invited to complete additional visits every 4 months.

### Measurements

A battery of instruments was assembled by a team of infectious disease clinicians and epidemiologists, aided by consultation with content specialists from pulmonology, cardiology, neurology, and mental health. Development of the instruments was an iterative process as information emerged regarding SARS-CoV-2 infection and recovery. Importantly, these instruments became adapted by other local studies of acute infection, facilitating a common measurement platform. These instruments queried about sociodemographic characteristics, medical history and concomitant medications, SARS-CoV-2 exposure, physical symptoms, quality of life, mental health, and substance use. With the exception of the mental health questions, which were self-administered, all questionnaires were interviewer-administered. Spanish translation was available.

At the initial visit, participants were asked to recount their physical symptoms and feelings during the period of acute infection as well as at the current moment. At all subsequent encounters, participants were asked about the time since the last visit and the present moment. The Patient Health Questionnaire (PHQ) somatic symptom scale [19] was used to ascertain physical symptoms. Participants were specifically asked to describe symptoms only if they were new or worse compared to the period prior to COVID-19. In addition to this predetermined list of symptoms, participants were asked about any other symptoms they were experiencing. Quality of life was measured using the EuroQol metrics [20], and mental health symptoms were measured using a combination of the General Anxiety Disorder-7 (GAD-7) [21], PHQ-8 [22], and an adaptive 4-item version of the post-traumatic stress disorder (PTSD) checklist (PCL) 5 [23–25]. In contrast to physical symptoms, questions about quality of life and mental health symptoms were not limited to new perceptions or feelings that occurred since onset of COVID-19; instead, they were answered from the perspective of the time of the interview.

### Statistical Analysis

Participants were described according to severity of COVID-19 during the first 21 days following onset of symptoms, which was classified as asymptomatic, symptomatic but not hospitalized, and hospitalized for the purposes of management of severe COVID-19. There were no hospitalizations solely for infection control or non-acute care. For each of the domains of physical symptoms, mental health symptoms and quality of life, we characterized 4 time periods: 1) acute illness (0-3 weeks), 2) early recovery (3-10 weeks), 3) late recovery 1 (12-20 weeks), and 4) late recovery 2 (28-36 weeks). We used Stata (version 16.1; StataCorp, College Station, TX) for these analyses.

## RESULTS

### Characteristics of Participants

From April 21, 2020 to January 4, 2021, we enrolled 179 adult participants who had experienced RNA-positivity-documented SARS-CoV-2 infection and were at least 14 days removed from the onset of symptoms, or if asymptomatic, the first detectable SARS-CoV-2 RNA. Most participants (60%) were enrolled within the first 3 months (April to July, 2020). At the time of enrollment, participants were a median of 1.8 months (interquartile range (IQR): 1.2 to 2.7) past the date of symptom onset/first positive RNA detection. The cohort represented the full spectrum of illness severity during the acute phase of SARS-CoV-2 infection (Table 1); 10 were asymptomatic, 125 were symptomatic but not hospitalized, and 44 were symptomatic and hospitalized. Among those who had been hospitalized, 37 (88%) required supplemental oxygen, but only 6 (14%) required mechanical ventilation. Few participants had received therapeutic interventions during acute SARS-CoV-2 infection; 6% received remdesivir, 10% glucocorticoids, and 2% convalescent plasma. The gender distribution (44% women) closely matched that of all cases in San Francisco (47% women; [26]). While there was diversity in race/ethnicity, those who self-identified as Hispanic/Latino (32% compared to 41% among all cases in San Francisco) and Asian (9.7% compared to 18% of all cases in San Francisco) were underrepresented and those who reported as White (51% compared to 22% of cases in San Francisco) were overrepresented. The majority of participants who had been hospitalized were men and of Hispanic/Latino ethnicity; non-hospitalized participants were more evenly distributed by gender and were predominantly White. Within the cohort, common medical comorbidities included hypertension, pre-COVID-19 lung disease, HIV infection, obesity, and diabetes. Thirty percent of participants had current or prior tobacco use.

**Table 1.**
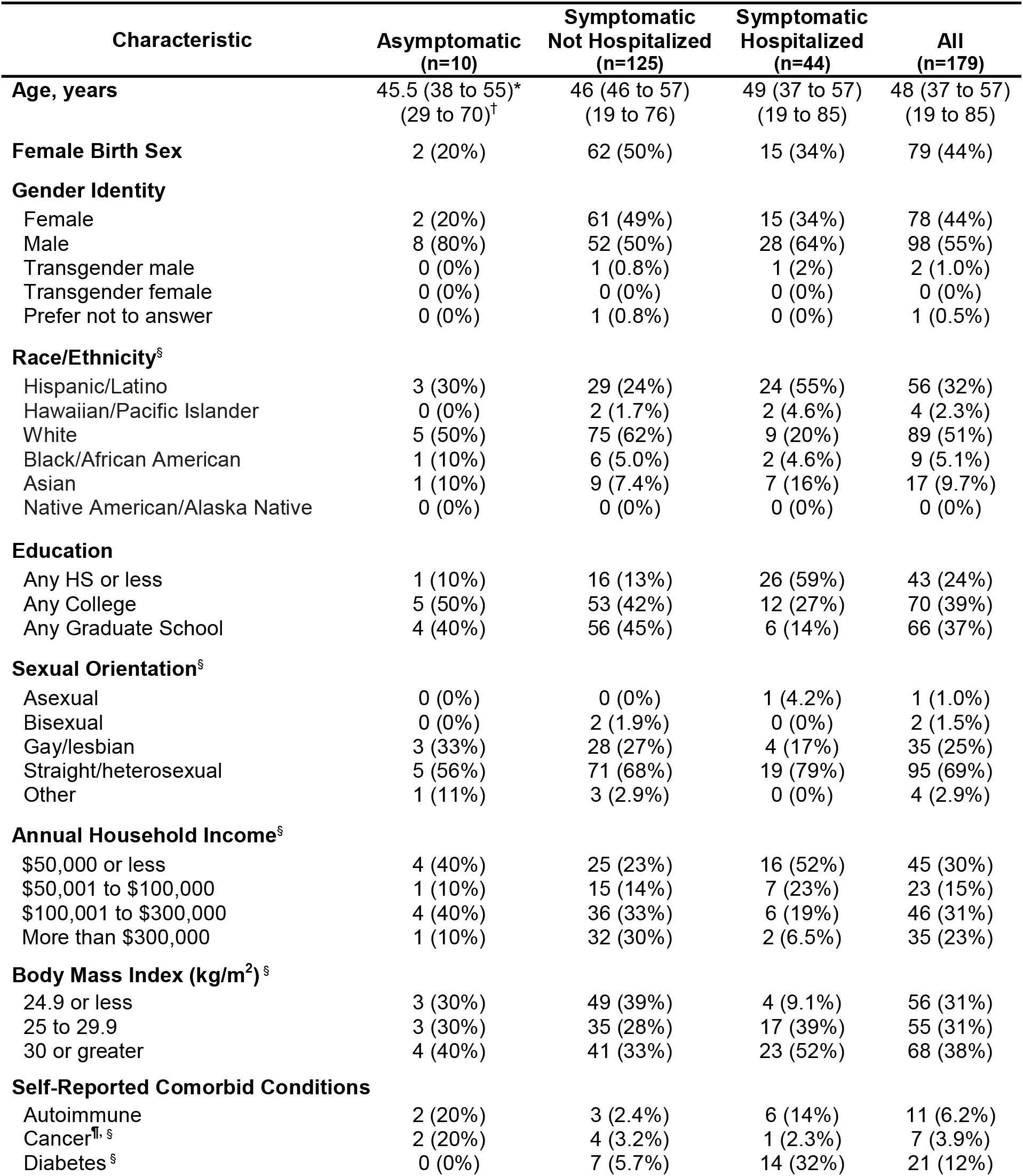
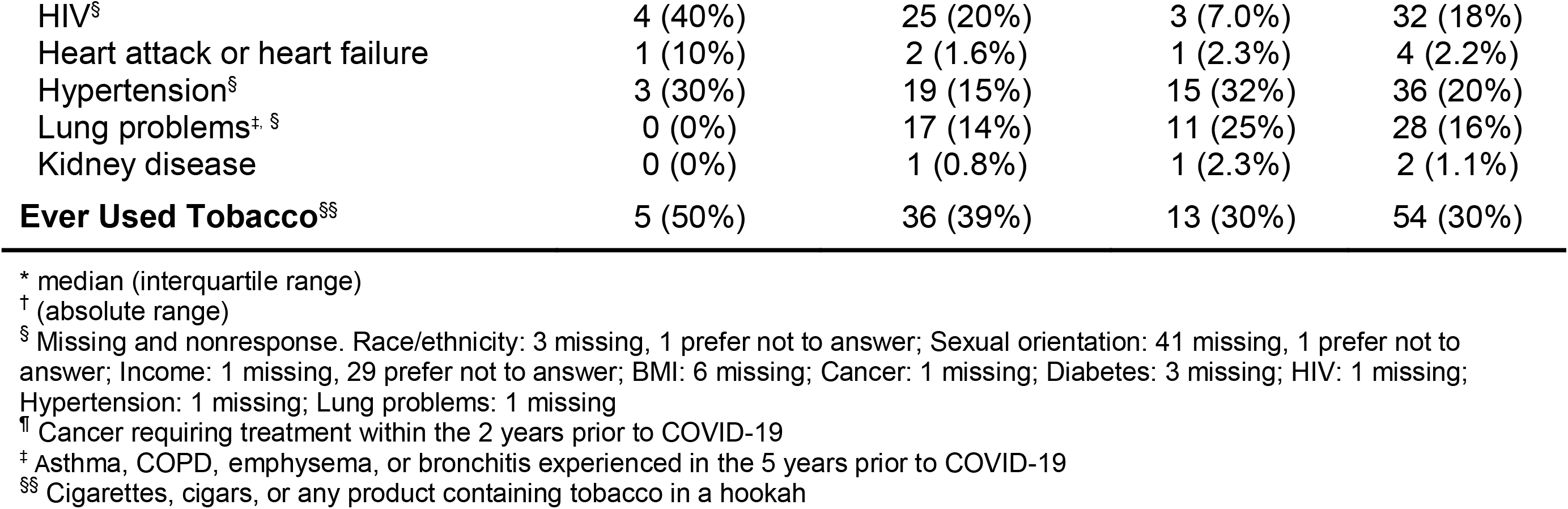
Characteristics at time of SARS-CoV-2 infection among participants enrolled in a study of post-acute sequelae of SARS-CoV-2 infection.

### Follow-up of Cohort

Longitudinal observation was scheduled every 4 months following onset of symptoms/date of first detected SARS-CoV-2 RNA. At 4 months, of the 165 participants who were evaluable (i.e., whose duration since symptom onset was at least 20.5 weeks, the outer boundary of the window for this visit), 143 (87%) completed the study visit. At 8 months, of the 111 participants who were evaluable, 68 (61%) completed the study visit. Of those who have not completed all follow-up visits, 10 have withdrawn from the study and none are known to have died.

### Physical Symptoms, Mental Health Symptoms, and Quality of Life

The most common symptoms during the acute phase of SARS-CoV-2 infection were fatigue, fever, myalgia, cough and anosmia/dysgeusia (Figure 1). In the post-acute phase time points, fatigue, shortness of breath, concentration problems, headaches, trouble sleeping and anosmia/dysgeusia were the most commonly reported, but a variety of other symptoms were endorsed by at least some participants at each time point. Importantly, these were not chronic symptoms, known to be present prior to COVID-19; they were instead symptoms that either newly developed or worsened since the onset of COVID-19. Not all participants, however, complained of symptoms. At early recovery (3-10 weeks), 61 participants reported no active symptoms. At the late recovery time points, 54 reported no active symptoms at the first late follow-up time point (12-20 weeks), and 16 reported no symptoms at the second late follow-up time point (28-36 weeks).

**Figure 1:**
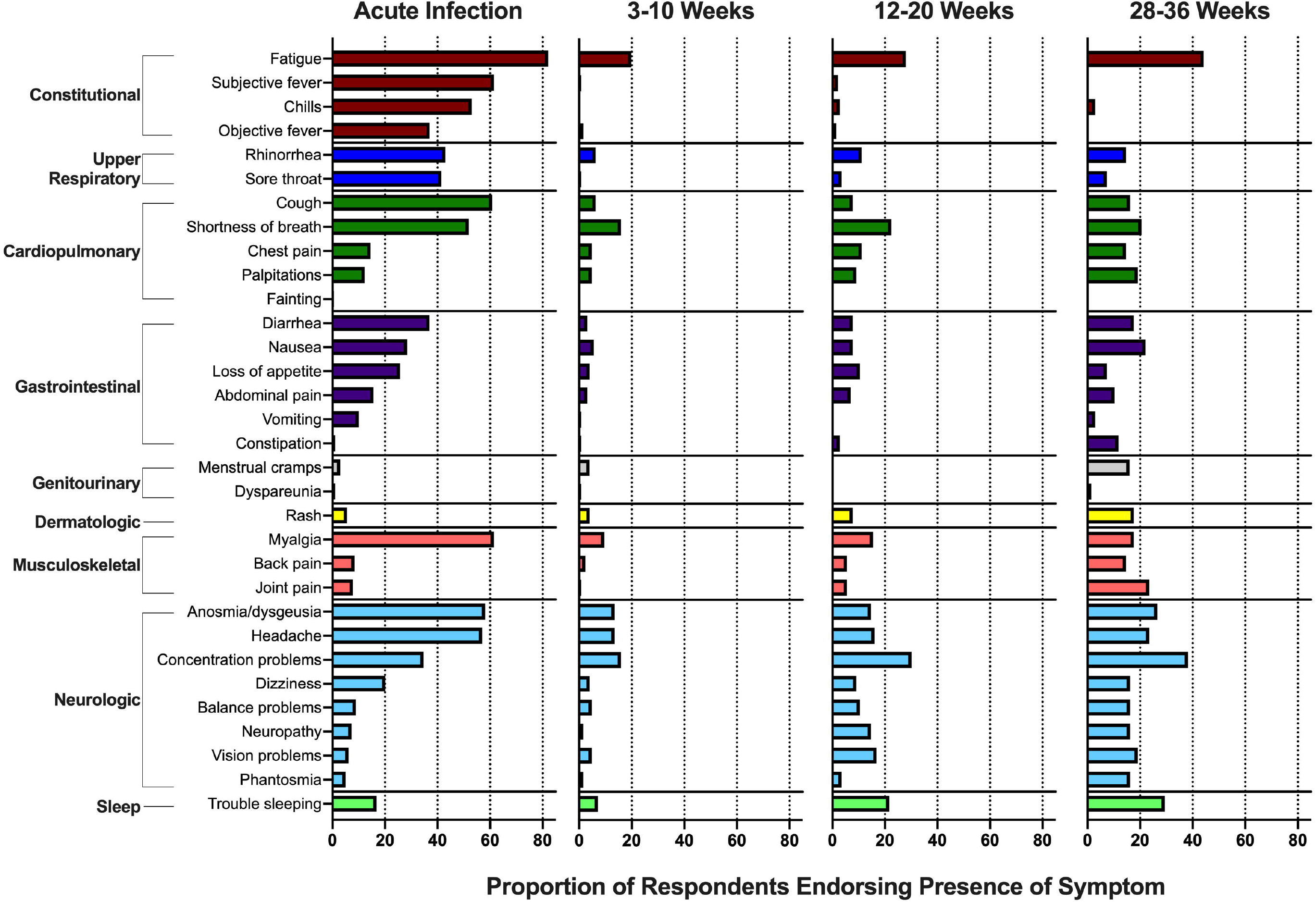
Prevalence of symptoms reported by participants in a study of individuals with SARS-CoV-2 infection during acute infection and three time points in the post-acute phase. Symptoms were limited to those not present prior to the occurrence of COVID-19. Concentration problems refers to “Trouble concentration, trouble with your thinking, or trouble with your memory.” Vision problems refers to “Trouble with vision, for example double vision, blurry vision, or other visual issues.”

Regarding mental health, some participants experienced symptoms of depression, anxiety and post-traumatic stress during the late recovery phase (Table 2). Most of these symptoms were described as minimal or mild, but a small number of individuals experienced moderate or severe symptoms. Measures of quality of life, which integrate across physical and mental health symptoms to depict functional impairments, showed expected high frequencies of inability to ambulate, perform self-care, and perform usual activities during the worst point of acute SARS-CoV-2 infection (Table 3). These frequencies were substantially higher than what participants reported prior to COVID-19. During the two late recovery periods, very few participants expressed moderate or more severe problems in self-care but some reported difficulties with ambulation and performance of usual activities. Participant-rated health on a visual analogue scale of 0 to 100 was, again, much lower during the worst point of acute infection compared to prior to COVID-19 (Table 3). In the two late recovery periods, the median visual analogue scale value was lower than prior to COVID-19.

**Table 2.** Responses regarding symptoms of anxiety, depression, and post-traumatic stress at 16 and 32 weeks following onset of COVID-19 among participants enrolled in a study of post-acute sequelae of SARS-CoV-2 infection.

| **Mental Health Symptoms and Severity** | **Week 16**<br>**(n = 119)\*** | **Week 32**<br>**(n = 65)\*** |
| --- | --- | --- |
| Symptoms of anxiety (GAD-7 total score)<sup>†</sup> |  |  |
| Minimal (0 to 4) | 68 (44%) | 33 (51%) |
| Mild (5 to 9) | 86 (56%) | 32 (49%) |
| Moderate (10 to 14) | 0 (0%) | 0 (0%) |
| Severe (15 to 21) | 0 (0%) | 0 (0%) |
| Symptoms of depression (PHQ-8 total score)<sup>†</sup> |  |  |
| None (0 to 4) | 70 (54%) | 39 (54%) |
| Mild (5 to 9) | 19 (17%) | 12 (20%) |
| Moderate (10 to 14) | 13 (12%) | 5 (8.0%) |
| Moderately severe (15 to 19) | 4 (3.5%) | 3 (5.0%) |
| Severe (20 to 24) | 4 (3.5%) | 2 (3.0%) |
| Symptoms of post-traumatic stress disorder (PCL-5 total score)<sup>†, §</sup> |  |  |
| Score ≥ 10 | 6 (6.0%) | 7 (11%) |

\* 24 responses at week 16 and 3 responses at week 32 are not expected due to differences in form versions used at the time of participant visit
<sup>†</sup> Missing and nonresponse. GAD-7: 2 missing at week 16; PHQ-8: 3 responses: 5 missing at week 16, missing at week 32; PCL-5: 8 missing at week 16, 1 missing at week 32.
<sup>§</sup> 4 item version of post-traumatic stress disorder checklist 5; cutoff of 10 has 76% sensitivity and 52% specificity for meeting the diagnostic criteria for post-traumatic stress disorder.

**Table 3.** Responses regarding quality of life prior to COVID-19, at the self-described worst point of COVID-19, and at weeks 16 and 32 following onset of COVID-19 among participants enrolled in a study of post-acute sequelae of SARS-CoV-2 infection.

| Quality of Life Domain | Response | Before Illness (n=92) | Worst Point (n=179) | Week 16 (n=117) | Week 32 (n=66) |
| --- | --- | --- | --- | --- | --- |
| **Mobility** | No problems | 91% | 39% | 85% | 77% |
|  | Slight problems | 6.0% | 10% | 9.0% | 11% |
| *“Which of the best describes your ability to walk about?”* | Moderate problems | 2.0% | 29% | 4.0% | 8.0% |
|  | Severe problems | 0% | 4.0% | 1.0% | 4.0% |
|  | Unable to walk | 1.0% | 18% | 0.0% | 0.0% |
| **Self-care** | No problems | 95% | 60% | 96% | 90% |
|  | Slight problems | 1.0% | 6.5% | 1.0% | 6.0% |
| *“Which of the following describes your ability to wash and dress yourself?”* | Moderate problems | 4.0% | 17% | 2.0% | 4.0% |
|  | Severe problems | 0% | 1.5% | 1.0% | 0% |
|  | Unable to wash or dress | 0% | 15% | 0% | 0% |
| **Usual activities** | No problems | 95% | 31% | 81% | 77% |
|  | Slight problems | 2.0% | 11% | 11% | 14% |
| *“Which of the following describes your ability to perform your usual activities?”* | Moderate problems | 1.0% | 23% | 6.0% | 1.0% |
|  | Severe problems | 2.0% | 7.0% | 2.0% | 5.0% |
|  | Unable to do usual activities | 0% | 28% | 0% | 3.0% |
| **Pain/discomfort\*** | No pain or discomfort | 70% | 27% | 65% | 52% |
|  | Slight pain or discomfort | 20% | 13% | 14% | 26% |
| *“Which of the following describes how much pain or discomfort you felt?”* | Moderate pain or discomfort | 7.0% | 26% | 17% | 15% |
|  | Severe pain or discomfort | 3.0% | 24% | 3.0% | 2.0% |
|  | Extreme pain or discomfort | 0% | 10% | 1.0% | 5.0% |
| **Anxiety/depression\*** | No anxiety or depression | 51% | 18% | 48% | 43% |
|  | Slight anxiety or depression | 27% | 23% | 31% | 32% |
| *“Which of the following describes how anxious or depressed you felt?”* | Moderate anxiety or depression | 17% | 24% | 15% | 14% |
|  | Severe anxiety or depression | 2.0% | 17% | 4.0% | 6.0% |
|  | Extreme anxiety or depression | 3.0% | 18% | 2.0% | 5.0% |
| **Visual analogue scale** |  |  |  |  |  |
|  | *“On a scale of 0 to 100, we would like to know how good or bad your health was...”* | 85<br>(75 to 90) | 50<br>(25 to 65) | 80<br>(70 to 90) | 80<br>(75 to 90) |

\* Not administered due to differences in form versions used at the time of participant visit. Pain/discomfort: 22 from week 16 and 1 from week 32; Anxiety/depression: 29 at week 16 and 1 from week 32

### Biologic Specimen Collection

A large number of biological specimens were collected from these participants and have been shared widely with internal and external investigators (Table 4). As of February 22, 2021, 1814 aliquots of PBMCs, 5316 aliquots of plasma, 3396 aliquots of serum and 545 gingival crevice salivary swabs have been distributed to research collaborators.

**Table 4.** Biological specimens collected and distributed as part of a study of post-acute

| Biological specimen | Number of participant-timepoints | Aliquots stored per participant-timepoint | Aliquots distributed |
| --- | --- | --- | --- |
| Cryopreserved PBMCs | 554 | 12 (6 to 28)* | 1814 |
| Plasma | 588 | 10 (9 to 29) | 5316 |
| Serum | 596 | 5 (4 to 14) | 3396 |
| Saliva, whole unstimulated | 210 | 10 (8 to 11) | 0 |
| Gingival crevice fluid | 596 | 1 (1 to 2) | 545 |

sequelae of SARS-CoV-2 infection.
\*median (IQR); PBMCs: peripheral blood mononuclear cells.

## DISCUSSION

As attention turns toward understanding the recovery phase following the acute manifestations of SARS-CoV-2 infection, large and well-characterized cohorts will be of great importance. Within two months of the first documented case of community transmission of SARS-CoV-2 in the U.S., we designed and initiated a cohort dedicated to studying the long-term impact of the infection. Despite ambient shelter-in-place conditions and other hardships suffered by the community, there has been substantial interest in the study and rapid enrollment. We have found that many people reported symptoms persisting beyond the acute illness period. These included generalized symptoms such as fatigue, organ system-specific manifestations such as cardiopulmonary and neurocognitive symptoms, and symptoms of anxiety and depression. While few participants endorsed major alterations in quality of life, some individuals have not resumed normal function during the recovery phase. With detailed phenotypic characterization and robust biologic specimen archiving, the cohort is now poised to support a variety of researchers to understand the causes of PASC.

Among a cohort of mostly non-hospitalized participants, we found a wide array of symptoms during the post-acute phase, many of which continued to be endorsed at the longest time point evaluable since acute illness. Our approach has extended upon earlier work about PASC and overcome several of the limitations of prior studies. Some of the largest reports to date have not required participants to have had confirmed SARS-CoV-2 infection [9], thus calling into question the specificity of the findings. We required documentation of SARS-CoV-2-RNA-positivity prior to enrollment. We included a large number of individuals who did not require hospitalization, and thus were able to be more representative of the entire spectrum of SARS-CoV-2 infection compared to other studies which focused on hospitalized patients [10,11]. We included up to 8 months of follow-up data, in contrast to studies which have been limited to the weeks and months following the initial illness [10,12,27]. Lastly, we have focused on biological specimen collection to facilitate biologically oriented research, which has already begun [28,29].

There are multiple mechanisms that might contribute to PASC, but the underlying mechanism or mechanisms have yet to be determined. It is likely that insults caused by the virus during the acute infectious period could initiate processes that lead to post-acute sequelae, even in the absence of ongoing antigen stimulation. However, one important consideration is whether viral antigen persists beyond the acute infectious period, either in the form of persistent virus replication [30] or persistence of non-infectious genetic material or protein in the tissues [31], where the virus has been demonstrated to be present in severe acute disease [32–35]. Regardless of whether the virus persists beyond acute infection, several mechanisms that are active in the recovery phase could explain PASC. First, systemic immune activation with alterations in B and T cell phenotypes and elevations in plasma cytokines and inflammatory markers could underlie at least some post-acute sequelae. These perturbations have been shown to be associated with more severe outcomes during acute infection [36,37]. Second, even in the absence of systemic inflammation, local tissue inflammation or ongoing immune cell infiltration into the tissues could result in tissue injury and remodeling, which could drive PASC through processes like microbial translocation in the gut [38] or tissue fibrosis in the heart or lungs. Organ-specific studies such as cardiac MRI suggest persistent inflammation for up to 3 months after acute illness [16,17]. Third, multiple studies, including autopsy studies, have demonstrated endotheliitis and microvascular thrombosis in acute COVID-19, with neutrophil extracellular traps as one contributing mechanism [39–44]; in addition to explaining severe disease, ongoing microvascular dysfunction may contribute to the pathobiology of PASC. Fourth, autoreactive immunity may be a significant contributor as IgG autoantibodies are highly prevalent in acute infection, including those associated with clinical disease entities similar to PASC [39,45–47]. Importantly, some mechanisms may contribute to certain organ-specific morbidity, whereas others might be common among various PASC phenotypes. Our cohort is well-positioned to ascertain which of these mechanisms are operative.

The nature of our participant sampling process limits what we can infer about PASC. Specifically, the urgency to study the long-term consequences of an infection that was affecting an already large and rapidly growing fraction of the population, the unprecedented workplace challenges and scarcity of resources forced us to decide to jumpstart this research by enrolling a convenience sample. It is axiomatic that our study population might be enriched for persons experiencing persistent symptoms because they were seeking answers for this condition. This may overestimate the parameter that all patients, clinicians and scientists wish to know, which is the prevalence (at, for example, 4 or 8 months) of persistent symptoms among all persons infected with SARS-CoV-2. Alternatively, it may be that persons who are most severely affected may be so debilitated that they were unable to travel to our research site. This would lead our study population to underestimate population-level prevalence. There is simply no means to know if our convenience sample is representative of the underlying target population as it relates to PASC, but there are suggestions that it is not. For example, empiric differences in our racial/ethnic distribution compared to all SARS-CoV-2 cases in San Francisco as well as a paucity of persons who had asymptomatic acute SARS-CoV-2 infection compared to what is believed to be a much higher fraction among all infected persons [48] suggest our population is not representative. It will only be through population-based probability samples that researchers can be confident that their study populations are representative of the relevant targets. For this reason, the percentage of study participants with persistent symptoms that we and others [9,49–51] calculate cannot be interpreted as meaningful population-level prevalence estimates. The threat of selection bias in the initial selection of participants also extends to our follow-up. We have experienced some losses to follow-up, and it may be that those participants with persistent symptoms are most motivated to continue in the study. The higher prevalence of some symptoms at 8 months compared to earlier time points that we observed might be a manifestation of this and thus should not be interpreted as symptoms getting worse over time.

Although our design precludes estimation of the population-level prevalence of PASC, there are several important aspects of PASC that it can investigate. First, conditioned upon the presence of persistent symptoms at the time of enrollment, our design can, if it maintains participant retention, yield relevant estimates of the duration of symptoms. Second, the presence of sizeable proportions of participants with and without ongoing symptoms during follow-up time points allows the cohort to serve as a resource (e.g., via efficient nested case-control designs) for investigating determinants of PASC. The detailed clinical characterization and biologic specimen collection performed at each recovery time point will allow us and interested collaborators to identify clinically relevant phenotypes and investigate the biologic causes of PASC.

In summary, we have established a cohort of participants enrolled in the post-acute phase of SARS-CoV-2 infection. We found substantial interest among affected community residents to participate, and among those who volunteered to enroll we found many who reported persistent physical symptoms through 8 months following onset of COVID-19. The convenience nature of our sampling — like many other nascent cohorts of PASC — precludes estimation of the population-level prevalence of these persistent symptoms, but it will allow for investigation of duration of symptoms conditioned upon their occurrence. Importantly, the presence of both participants without symptoms and ample biological specimens will also allow for analytic work to study the pathogenesis of PASC, which we anticipate will burgeon as the field begins to investigate mechanisms that will inform prevention and treatment.

## Data Availability

This is an ongoing study. Data are available upon request and will be provided at the discretion of the study leadership team.

## ACKNOWLEDGEMENTS

We are grateful to the LIINC study participants and to the clinical staff who provided care to these individuals during their acute illness period and during their recovery. We acknowledge LIINC clinical study team members Tamara Abualhsan, Heather Hartig, Marian Kerbleski, Cassandra Thanh, and Fatima Ticas; LIINC laboratory team members Joanna Donatelli, Jill Hakim, Nikita Iyer, Owen Janson, Christopher Nixon, and Keirstinne Turcios; and medical student volunteers Isabella Auchus, T. Allen Barnett, and Chloe Cattle. We thank Elnaz Eilkhani for coordination with the Institutional Review Board. We are grateful to LIINC collaborators Fran Aweeka, Jessica Briggs, Felicia Chow, Amelia Deitchman, Meredith Greene, Joanna Hellmuth, Laurence Huang, Peter Hunt, Mandana Khalili, Sulggi Lee, Lynn Pulliam, Edda Santiago Rodriguez, Lekshmi Santhosh, Ma Somsouk, and Joshua Vasquez for their scientific advice and collaboration. We acknowledge the contributions of the UCSF Clinical and Translational Science Unit, Core Immunology Laboratory, and AIDS Specimen Bank, as well as support and advice from the Division of HIV, Infectious Diseases, and Global Medicine and ZSFG Infection Control Service, in particular Diane Havlir, Vivek Jain, Carina Marquez, Lisa Winston, and Annie Luetkemeyer.

## AUTHOR CONTRIBUTIONS

MJP, JDK, TJH, SGD, and JNM designed the cohort, which was overseen by MJP, JDK, RLR, IRB, BG, TJH, SGD, and JNM. MJP, JDK, SL, MCD, RH, HMS, SGD, and JNM designed the study instruments with input from VT, EAF, LT, YH, and KA; EOM, KAA, and JP-R assisted with Spanish translation. MJP, RH, VT, EAF, LT, YH, MIW, MIA, JAB, LHN, MD, SEM and EMO recruited participants and coordinated research visits. MJP, JDK, MCD, RH, VT, EAF, LT, YH, MIA, JAB, LHN, and SEM administered study questionnaires and collected clinical data. JAB, MCW, and MDR collected biospecimens. SL, SAG, MCD, SM, MD, EOM, KAA, MDR, JT, PRR, HMS, VWM, PKE, KCD, JAM, JOW, IM, JP-R, and AFT performed data entry and validation. MW, MA, YY, MK, and JNM designed and maintained the study database. SL, SAG, MDC, SM cleaned the data and performed the analyses, which were planned by MJP, JDK, and JNM. MJP, JDK, MSD, and JNM drafted the manuscript, with extensive input from MAS, RLR, IRB, BG, JAS, MG, PYH, SGD, and TJH. The study was primarily funded by MAS, MG, SGD, and TJH with additional support from MJP, JDK, BG, PYH, and JNM. All authors reviewed, edited, and approved the manuscript.

## REFERENCES

1 Guan W-J, Ni Z-Y, Hu Y, Liang W-H, Ou C-Q, He J-X, et al. Clinical Characteristics of Coronavirus Disease 2019 in China. N Engl J Med 2020; 382:1708–1720.

2 Goyal P, Choi JJ, Pinheiro LC, Schenck EJ, Chen R, Jabri A, et al. Clinical Characteristics of Covid-19 in New York City. N Engl J Med 2020; 382:2372–2374.

3 Richardson S, Hirsch JS, Narasimhan M, Crawford JM, McGinn T, Davidson KW, et al. Presenting Characteristics, Comorbidities, and Outcomes Among 5700 Patients Hospitalized With COVID-19 in the New York City Area. JAMA 2020; 323:2052–2059.

4 Harding L. “Weird as hell”: the Covid-19 patients who have symptoms for months. The Guardian. 2020.http://www.theguardian.com/world/2020/may/15/weird-hell-professor-advent-calendar-covid-19-symptoms-paul-garner (accessed 7 Mar2021).

5 Chuck E, Edwards E. Doctors couldn’t help these COVID-19 patients with their endless symptoms. So they turned to one another. NBC News. 2020.https://www.nbcnews.com/health/health-news/doctors-couldn-t-help-these-covid-19-patients-their-endless-n1208116 (accessed 7 Mar2021).

6 Horowitz J. Surviving Covid-19 May Not Feel Like Recovery for Some. The New York Times. 2020.https://www.nytimes.com/2020/05/10/world/europe/coronavirus-italy-recovery.html (accessed 7 Mar2021).

7 Yong E. COVID-19 Can Last for Several Months. The Atlantic 2020.https://www.theatlantic.com/health/archive/2020/06/covid-19-coronavirus-longterm-symptoms-months/612679/ (accessed 7 Mar2021).

8 Yong E. Long-Haulers Are Redefining COVID-19. The Atlantic 2020.https://www.theatlantic.com/health/archive/2020/08/long-haulers-covid-19-recognition-support-groups-symptoms/615382/ (accessed 7 Mar2021).

9 Davis HE, Assaf GS, McCorkell L, Wei H, Low RJ, Re’em Y, et al. Characterizing Long COVID in an International Cohort: 7 Months of Symptoms and Their Impact. medRxiv Published Online First: 2020.https://www.medrxiv.org/content/10.1101/2020.12.24.20248802v1.full-text

10 Carfì A, Bernabei R, Landi F, Gemelli Against COVID-19 Post-Acute Care Study Group. Persistent Symptoms in Patients After Acute COVID-19. JAMA 2020; 324:603–605.

11 Huang C, Huang L, Wang Y, Li X, Ren L, Gu X, et al. 6-month consequences of COVID-19 in patients discharged from hospital: a cohort study. Lancet 2021; 397:220–232.

12 Tenforde MW, Kim SS, Lindsell CJ, Billig Rose E, Shapiro NI, Files DC, et al. Symptom Duration and Risk Factors for Delayed Return to Usual Health Among Outpatients with COVID-19 in a Multistate Health Care Systems Network - United States, March-June 2020. MMWR Morb Mortal Wkly Rep 2020; 69:993–998.

13 Liu D, Zhang W, Pan F, Li L, Yang L, Zheng D, et al. The pulmonary sequalae in discharged patients with COVID-19: a short-term observational study. Respir Res 2020; 21:125.

14 Liu C, Ye L, Xia R, Zheng X, Yuan C, Wang Z, et al. Chest Computed Tomography and Clinical Follow-Up of Discharged Patients with COVID-19 in Wenzhou City, Zhejiang, China. Ann Am Thorac Soc 2020; 17:1231–1237.

15 Zhao Y-M, Shang Y-M, Song W-B, Li Q-Q, Xie H, Xu Q-F, et al. Follow-up study of the pulmonary function and related physiological characteristics of COVID-19 survivors three months after recovery. EClinicalMedicine 2020; 25:100463.

16 Puntmann VO, Carerj ML, Wieters I, Fahim M, Arendt C, Hoffmann J, et al. Outcomes of Cardiovascular Magnetic Resonance Imaging in Patients Recently Recovered From Coronavirus Disease 2019 (COVID-19). JAMA Cardiol 2020; 5:1265–1273.

17 Huang L, Zhao P, Tang D, Zhu T, Han R, Zhan C, et al. Cardiac Involvement in Patients Recovered From COVID-2019 Identified Using Magnetic Resonance Imaging. JACC Cardiovasc Imaging 2020; 13:2330– 2339.

18 Bonny TS, Patel EU, Zhu X, Bloch EM, Grabowski MK, Abraham AG, et al. Cytokine and Chemokine Levels in Coronavirus Disease 2019 Convalescent Plasma. Open Forum Infect Dis 2021; 8:ofaa574.

19 Kroenke K, Spitzer RL, Williams JBW. The PHQ-15: validity of a new measure for evaluating the severity of somatic symptoms. Psychosom Med 2002; 64:258–266.

20 Rabin R, Charro F de. EQ-SD: a measure of health status from the EuroQol Group. Ann Med 2001; 33:337–343.

21 Spitzer RL, Kroenke K, Williams JBW, Löwe B. A brief measure for assessing generalized anxiety disorder: the GAD-7. Arch Intern Med 2006; 166:1092–1097.

22 Kroenke K, Strine TW, Spitzer RL, Williams JBW, Berry JT, Mokdad AH. The PHQ-8 as a measure of current depression in the general population. J Affect Disord 2009; 114:163–173.

23 Cameron RP, Gusman D. The primary care PTSD screen (PC-PTSD): development and operating characteristics. Primary care psychiatry 2003; 9:9–14.

24 Blevins CA, Weathers FW, Davis MT, Witte TK, Domino JL. The Posttraumatic Stress Disorder Checklist for DSM-5 (PCL-5): Development and initial psychometric evaluation: Posttraumatic stress disorder checklist for DSM-5. J Trauma Stress 2015; 28:489–498.

25 Price M, Szafranski DD, van Stolk-Cooke K, Gros DF. Investigation of abbreviated 4 and 8 item versions of the PTSD Checklist 5. Psychiatry Res 2016; 239:124–130.

26 COVID-19 Data and Reports. https://data.sfgov.org/stories/s/fjki-2fab (accessed 7 Mar2021).

27 Carvalho-Schneider C, Laurent E, Lemaignen A, Beaufils E, Bourbao-Tournois C, Laribi S, et al. Follow-up of adults with noncritical COVID-19 two months after symptom onset. Clin Microbiol Infect Published Online First: 5 October 2020. doi:10.1016/j.cmi.2020.09.052

28 Peluso MJ, Takahashi S, Hakim J, Kelly JD, Torres L, Iyer NS, et al. SARS-CoV-2 antibody magnitude and detectability are driven by disease severity, timing, and assay. medRxiv Published Online First: 5 March 2021. doi:10.1101/2021.03.03.21251639

29 Peluso MJ, Deitchman AN, Torres L, Iyer NS, Nixon CC, Munter SE, et al. Long-Term SARS-CoV-2-Specific Immune and Inflammatory Responses Across a Clinically Diverse Cohort of Individuals Recovering from COVID-19. medRxiv Published Online First: 1 March 2021. doi:10.1101/2021.02.26.21252308

30 Zheng S, Fan J, Yu F, Feng B, Lou B, Zou Q, et al. Viral load dynamics and disease severity in patients infected with SARS-CoV-2 in Zhejiang province, China, January-March 2020: retrospective cohort study. BMJ 2020; 369:m1443.

31 Gaebler C, Wang Z, Lorenzi JCC, Muecksch F, Finkin S, Tokuyama M, et al. Evolution of antibody immunity to SARS-CoV-2. Nature Published Online First: 18 January 2021. doi:10.1038/s41586-021-03207-w

32 Remmelink M, De Mendonça R, D’Haene N, De Clercq S, Verocq C, Lebrun L, et al. Unspecific post-mortem findings despite multiorgan viral spread in COVID-19 patients. Crit Care 2020; 24:495.

33 Song E, Zhang C, Israelow B, Lu-Culligan A, Prado AV, Skriabine S, et al. Neuroinvasion of SARS-CoV-2 in human and mouse brain. J Exp Med 2021; 218. doi:10.1084/jem.20202135

34 Lindner D, Fitzek A, Bräuninger H, Aleshcheva G, Edler C, Meissner K, et al. Association of Cardiac Infection With SARS-CoV-2 in Confirmed COVID-19 Autopsy Cases. JAMA Cardiol 2020; 5:1281–1285.

35 Bussani R, Schneider E, Zentilin L, Collesi C, Ali H, Braga L, et al. Persistence of viral RNA, pneumocyte syncytia and thrombosis are hallmarks of advanced COVID-19 pathology. EBioMedicine 2020; 61:103104.

36 Lucas C, Wong P, Klein J, Castro TBR, Silva J, Sundaram M, et al. Longitudinal analyses reveal immunological misfiring in severe COVID-19. Nature 2020; 584:463–469.

37 Laing AG, Lorenc A, del Molino del Barrio I, Das A, Fish M, Monin L, et al. A dynamic COVID-19 immune signature includes associations with poor prognosis. Nat Med 2020; 26:1623–1635.

38 Giron LB, Dweep H, Yin X, Wang H, Damra M, Goldman AR, et al. Severe COVID-19 is fueled by disrupted gut barrier integrity. bioRxiv. 2020. doi:10.1101/2020.11.13.20231209

39 Zuo Y, Estes SK, Ali RA, Gandhi AA, Yalavarthi S, Shi H, et al. Prothrombotic autoantibodies in serum from patients hospitalized with COVID-19. Sci Transl Med 2020; 12. doi:10.1126/scitranslmed.abd3876

40 Gu SX, Tyagi T, Jain K, Gu VW, Lee SH. Thrombocytopathy and endotheliopathy: crucial contributors to COVID-19 thromboinflammation. Nat Rev Published Online First: 2020.https://www.nature.com/articles/s41569-020-00469-1?elqTrackId=1463b6ba0cd94325a79ebd0e5db4310e

41 Goshua G, Pine AB, Meizlish ML, Chang C-H, Zhang H, Bahel P, et al. Endotheliopathy in COVID-19-associated coagulopathy: evidence from a single-centre, cross-sectional study. The Lancet Haematology 2020; 7:e575–e582.

42 Rauch Antoine, Dupont Annabelle, Goutay Julien, Caplan Morgan, Staessens Senna, Moussa Mouhamed, et al. Endotheliopathy Is Induced by Plasma From Critically Ill Patients and Associated With Organ Failure in Severe COVID-19. Circulation 2020; 142:1881–1884.

43 Maccio U, Zinkernagel AS, Shambat SM, Zeng X, Cathomas G, Ruschitzka F, et al. SARS-CoV-2 leads to a small vessel endotheliitis in the heart. EBioMedicine 2021; 63:103182.

44 Varga Z, Flammer AJ, Steiger P, Haberecker M, Andermatt R, Zinkernagel AS, et al. Endothelial cell infection and endotheliitis in COVID-19. The Lancet. 2020; 395:1417–1418.

45 Chang SE, Feng A, Meng W, Apostolidis SA, Mack E, Artandi M, et al. New-onset IgG autoantibodies in hospitalized patients with COVID-19. medRxiv Published Online First: 29 January 2021. doi:10.1101/2021.01.27.21250559

46 Zhou Y, Han T, Chen J, Hou C, Hua L, He S, et al. Clinical and Autoimmune Characteristics of Severe and Critical Cases of COVID-19. Clin Transl Sci 2020; 13:1077–1086.

47 Bhadelia N, Belkina AC, Olson A, Winters T, Urick P, Lin N, et al. Distinct autoimmune antibody signatures between hospitalized acute COVID-19 patients, SARS-CoV-2 convalescent individuals, and unexposed pre-pandemic controls. bioRxiv. 2021. doi:10.1101/2021.01.21.21249176

48 Yanes-Lane M, Winters N, Fregonese F, Bastos M, Perlman-Arrow S, Campbell JR, et al. Proportion of asymptomatic infection among COVID-19 positive persons and their transmission potential: A systematic review and meta-analysis. PLoS One 2020; 15:e0241536.

49 Logue JK, Franko NM, McCulloch DJ, McDonald D, Magedson A, Wolf CR, et al. Sequelae in Adults at 6 Months After COVID-19 Infection. JAMA Netw Open 2021; 4:e210830.

50 Jacobson KB, Rao M, Bonilla H, Subramanian A, Hack I, Madrigal M, et al. Patients with uncomplicated COVID-19 have long-term persistent symptoms and functional impairment similar to patients with severe COVID-19: a cautionary tale during a global pandemic. Clin Infect Dis Published Online First: 7 February 2021. doi:10.1093/cid/ciab103

51 Hellmuth J, Allen Barnett T, Asken BM, Daniel Kelly J, Torres L, Stephens ML, et al. Persistent COVID-19-associated neurocognitive symptoms in non-hospitalized patients. Journal of NeuroVirology. 2021. doi:10.1007/s13365-021-00954-4

